# Enhanced Conduit Flow Compensates for the Reduction in Left Atrial Passive and Booster Functions in Advanced Diastolic Dysfunction

**DOI:** 10.1101/2023.08.01.23293445

**Authors:** Doron Aronson, Hend Sliman, Sobhi Abadi, Ida Maiorov, Daniel Perlow, Diab Mutlak, Jonathan Lessick

**Author notes:** Address for Correspondence: Doron Aronson, MD Department of Cardiology Rambam Medical Center POB 9602, Haifa 31096, Israel.

## Abstract

**Background:** Quantification of left atrial (LA) conduit function and its contribution to left ventricular (LV) filling is challenging because it requires simultaneous measurements of both LA and LV volumes. The functional relationship between LA conduit function and the severity diastolic dysfunction remains controversial. We studied the role of LA conduit function in maintaining LV filling in advanced diastolic dysfunction.

**Methods:** We performed volumetric and flow analyses of LA function across the spectrum of LV diastolic dysfunction, derived from a set of consecutive patients undergoing multiphasic cardiac CT scanning (n=489). From LA and LV time-volume curves we calculated 3 volumetric components: 1) early “passive” emptying volume; 2) late “active” (booster) volume; and 3) conduit volume. Results were prospectively validated on a group of patients with severe aortic stenosis (n=110).

**Results:** The early passive filling progressively decreased with worsening diastolic function (*P*<0.0001). The atrial booster contribution to stroke volume (SV) modestly increases in impaired relaxation (*P*<0.05) and declined with more advanced diastolic function (*P*<0.001), thus failing to compensate for the reduction in early filling. The conduit volume increased progressively (*P*<0.0001), accounting for 75% of SV (IQR 63–81%) with restrictive filling pattern, compensating for the reduction in both early and booster functions. Similar results were obtained in patients with severe aortic stenosis. The pulmonary artery systolic pressure increased in a near-linear fashion when the conduit contribution to SV increased above 60%. Maximal conduit flow rate strongly correlated with mitral E-wave velocity (r=0.71, *P*<0.0001), indicating that the increase in mitral E-wave in diastolic dysfunction represents the increased conduit flow.

**Conclusion:** An increase in conduit volume contribution to SV represents a compensatory mechanism to maintain LV filling in advanced diastolic dysfunction. The increase in conduit volume despite increasing LV diastolic pressures is accomplished by an increase in pulmonary venous pressure.

## Introduction

The left atrium (LA) plays an important role in the maintenance of cardiovascular homeostasis and assumes greater importance in heart failure (HF) (1). Pathological conditions resulting in elevation of LA pressure trigger atrial structural and functional changes (atrial remodeling) characterized by LA dilatation and impaired LA functional parameters (2,3). LA remodeling correlates with the severity of diastolic dysfunction and contributes to symptoms (4,5) and to disease progression (2).

The contribution of LA function to left ventricular (LV) filling, and hence LV stroke volume (SV), involves 3 components: 1) in early diastole, blood stored in LA is driven into the LV (LA passive emptying) predominantly via the atrioventricular pressure gradient developed by LV suction and partially by elastic energy stored by the LA during the reservoir phase (6); 2) Conduit flow, that involves the transfer of blood into the LV from the pulmonary veins during early LV filling and diastasis without affecting LA volume (i.e., volume of blood that passes through the LA that cannot be accounted for by reservoir or booster pump function (7); 3) Active atrial contraction, also referred as LA booster pump function (1).

The current paradigm of diastolic dysfunction focuses on progressive impairment of LV relaxation and compliance and subsequent atrial failure with no reference to the role of conduit flow (3,8). The relationship between conduit volume and the severity of diastolic dysfunction remains controversial. The majority of studies suggest that LA conduit function progressively decreases with aging and increasing grades of LV diastolic dysfunction (LVDD), with a compensatory increase in booster function to maintain total LA emptying volumes (9–11). Other studies have found that conduit function increases with advanced stages (12). This is partly derived from the current confusion in the literature regarding the definition of “conduit function”.

Quantification of LA conduit function is challenging because it requires simultaneous measurements of the LA and LV volume changes (13). Therefore, passive emptying and conduit are often combined into a single “conduit” phase (14,15). In the present study we employed cardiac computed tomography angiography CTCA to acquire complete and simultaneous LA and LV volumetric data to characterize the 3 components of LV diastolic filling (16). We show that LA conduit function plays a key role in maintaining LV filling in advanced LVDD and strongly correlates with the development of pulmonary hypertension.

## Methods

The study was approved by the local institutional review board. From our database of patients who had undergone CCTA, we identified patients who had undergone both a cardiac CT examination, using spiral scanning with retrospective gating, as well as a full transthoracic echocardiogram (TTE) within a period of 30-days prior or after the day of the index CT. Scanning was performed on a dual-source Somatom Definition Flash scanner (Siemens Healthcare). Data was reconstructed every 5% of the cardiac cycle to provide 20 phases per study. We excluded patients with chronic atrial fibrillation, unstable patients, insufficient CT or echo quality, and indeterminate LVDD grade. In addition, we prospectively validated the results of the retrospective analysis on patients with severe aortic stenosis (AS) referred for TAVI.

Data were analyzed on a dedicated CT workstation (IntelliSpace Portal, version 8, Philips Healthcare). Retrospective data were analyzed using fully automatic segmentation of the heart chambers, producing phasic volume curves of each heart chamber. LA volume–based indices were calculated as previously described (17,18). In the AS group, manual editing was used to enhance the accuracy of the measurements (16).

For each patient, calculated time-volumes curves were used to identify LV end-diastolic volume (LVEDV), LV end-systolic volume (LVESV), LA maximum end-systolic volume (LAESV), LA pre-A volume and LA minimum end-diastolic volume (LAEDV) as well as LV mass (LVM) at end-diastole. LV ejection fraction was calculated as LVEF=100*(LVEDV-LVESV)/LVEDV.

Total emptying function (LAEF**_Total_**), passive LAEF (LAEF**_Passive_**) and active LAEF (LAEF**_Contractile_**) were defined as fractional volume changes as follows: LAEF**_total_** = (LAV**_max_**- LAV**_min_**)/ LAV**_max_**, LAEF**_Passive_** = (LAV**_max_**-LAV**_pre-a_**)/LAV**_max_** and LAEF**_Contractile_**=(LAV**_pre-a_** – LAVmin)/LAV**_pre-a_** (19,20). LA expansion index, representing the relative LA volume changes during the reservoir phase, was calculated as LAEI = (LAV**_max_** - LAV**_min_**)/LAV**_min_**) (15,16,18,21–23).

Based on these calculations, LV filling was divided into 3 volumetric components : 1) early “passive” emptying volume; 2) late “active” (booster) volume; and 3) conduit volume, defined as the volume of blood that passes through the LA that cannot be accounted for by early or booster pump function: LV stroke volume - (LA**_max_** volume – LA**_min_** volume)] (2). These components of the LA function were expressed in terms of LV filling as percent contribution to left ventricular SV (7).

In a subset of 55 patients, conduit volume was calculated for each cardiac phase (t) from the start of diastole (t=0) to the atrial kick. This was done using the formula: LA conduit volume (t) = [LV(t) – LVESV] – [LAESV – LA(t)] for t=0 to t=phase at start of atrial kick (7). The duration of each phase is ∼50msec, depending on the patient’s heart rate. This produces a positive slope for conduit volume from 0 to maximum. The maximal conduit flow rate was also calculated and correlated with the echocardiographic mitral E wave.

A full echocardiographic diastolic assessment was carried out according to the ASE recommendations (8), as previously described (21). Two experienced cardiologists reviewed all TTE reports for assessment of diastolic function, and classified them as normal, grade I-III LVDD, or indeterminate, based on ASE recommendations (8). Patients with indeterminate diastolic function were excluded.

### Statistical analysis

Continuous variables are presented as mean±SD or medians (with interquartile ranges), and categorical variables as numbers and percentages. The relationship between basic parameters measured by echo and by CT, were assessed by Pearson correlation. The relationship between echocardiographic grading of diastolic function and the 3 volumetric components of LA function were analyzed using the Kruskal–Wallis test followed by post-hoc Dunn’s test with the Holm-Sidák correction for multiple comparisons.

The rate of conduit flow (measured from start of diastole to the start of the atrial kick) into the LV was analyzed using a linear mixed-model with random intercepts. The model included an indicator variable for diastolic function, time, and an interaction term between diastolic function and time. The time was modeled using a quadratic term to allow for some curvature in the relationship.

The relation between the contribution of conduit volume to SV and pulmonary artery systolic pressure was estimated with the use of restricted cubic splines (with 3 knots placed at default locations), which allowed us to explore nonlinear relationships. Differences were considered statistically significant at the 2-sided P<0.05 level. Statistical analyses were performed using STATA Version 17.0 (College Station, TX).

## Results

The study cohort comprised of 1067 patients in sinus rhythm who had undergone spiral CTCA at our institution between January 2010 to June 2018. After exclusion of patients with no recent echocardiography exam, chronic atrial fibrillation, insufficient CT or echo quality, and patients with indeterminate LVDD grade, 489 patients remained. The AS group included 183 patients undergoing spiral CTCA from January 2017 to December 2021. After similar exclusions, 110 patients remained for analysis. The clinical characteristics and CCTA finding of the 2 study groups are summarized in Table 1.

**Table 1:** Baseline Characteristics

| **Characteristics** | **Retrospective**<br>**(n = 489)** | **Aortic Stenosis**<br>**(n = 110)** |
| --- | --- | --- |
| Age (years) | 67 ± 15 | 78 ± 10 |
| Male gender | 296 (61) | 54 (49) |
| Body surface area (m<sup>2</sup>) | 1.89 ± 0.22 | 1.84 ± 0.23 |
| HTN | 262 (74) | 79 (88) |
| DM | 180 (37) | 48 (44) |
| Coronary artery disease | 214 (44) | 77 (70) |
| Previous CABG | 94 (19) | 12 (11) |
| Previous MI | 134 (27) | 41 (37) |
| Previous HF | 94 (19) | 32 (29) |
| **Echocardiographic diastolic grade** |  |  |
| Normal | 133 (27) | 2 (2) |
| Impaired relaxation | 88 (18) | 19 (17) |
| Pseudonormal | 215 (44) | 78 (71) |
| Restrictive pattern | 53 (11) | 11 (10) |
| **CT Measurements** |  |  |
| LVEDVI (mL/m<sup>2</sup>) | 91 ± 21 | 91 ± 25 |
| LVESVI (mL/m<sup>2</sup>) | 39 ± 24 | 44 ± 22 |
| LVEF (%) | 60 ± 14 | 54 ± 13 |
| LVMI (g/m<sup>2</sup>) | 84 ± 28 | 87 ± 26 |
| LAESVI (ml/m<sup>2</sup>) | 61 ± 17 | 65 ± 14 |
| LAEDVI (ml/m<sup>2</sup>) | 39 ± 17 | 44 ± 14 |
| LAV<sub>pre-A</sub> (mL) | 98 ± 30 | 103 ± 25 |
| LAEF<sub>Passive</sub> (%) | 15 ± 7 | 12 ± 4 |
| LAEF<sub>Contractile</sub> (%) | 22 ± 9 | 24 ± 11 |
| LAEF<sub>Total</sub> (%) | 39 ± 12 | 33 ± 11 |

Values are presented as n (%), mean ± SD or median (interquartile range).

### Relationship between phasic changes in LA volume and echocardiographic diastolic function

The changes in the early filling, booster, and conduit volume contributions to SV according to diastolic function are depicted in Figure 1. The early passive filling component was highest in patients with normal diastolic function and progressively decreased as diastolic function worsened (*P*=0.01, *P*<0.0001 and *P*<0.0001 for the comparison of impaired relaxation, pseudonormal and restrictive filling with normal diastolic function, respectively). The atrial booster contribution to SV modestly increases in patients with impaired relaxation compared with those with normal diastolic function (*P*<0.05) but declines with the more advance diastolic function grades and becomes significantly lower than normal values in restrictive filling pattern (*P*<0.001). Hence, in patients with impaired relaxation, the increase in booster function compensates for the decline in early diastolic filling but with grade II diastolic dysfunction (pseudonormal filling), the booster contribution to SV returns to its original value and cannot compensate for the reduction in early passive filling that continues to decline. In grade III diastolic dysfunction (restrictive filling pattern), the contribution of both early and booster functions to SV was only 25% [IQR 20 to 35%].

**Figure 1:**
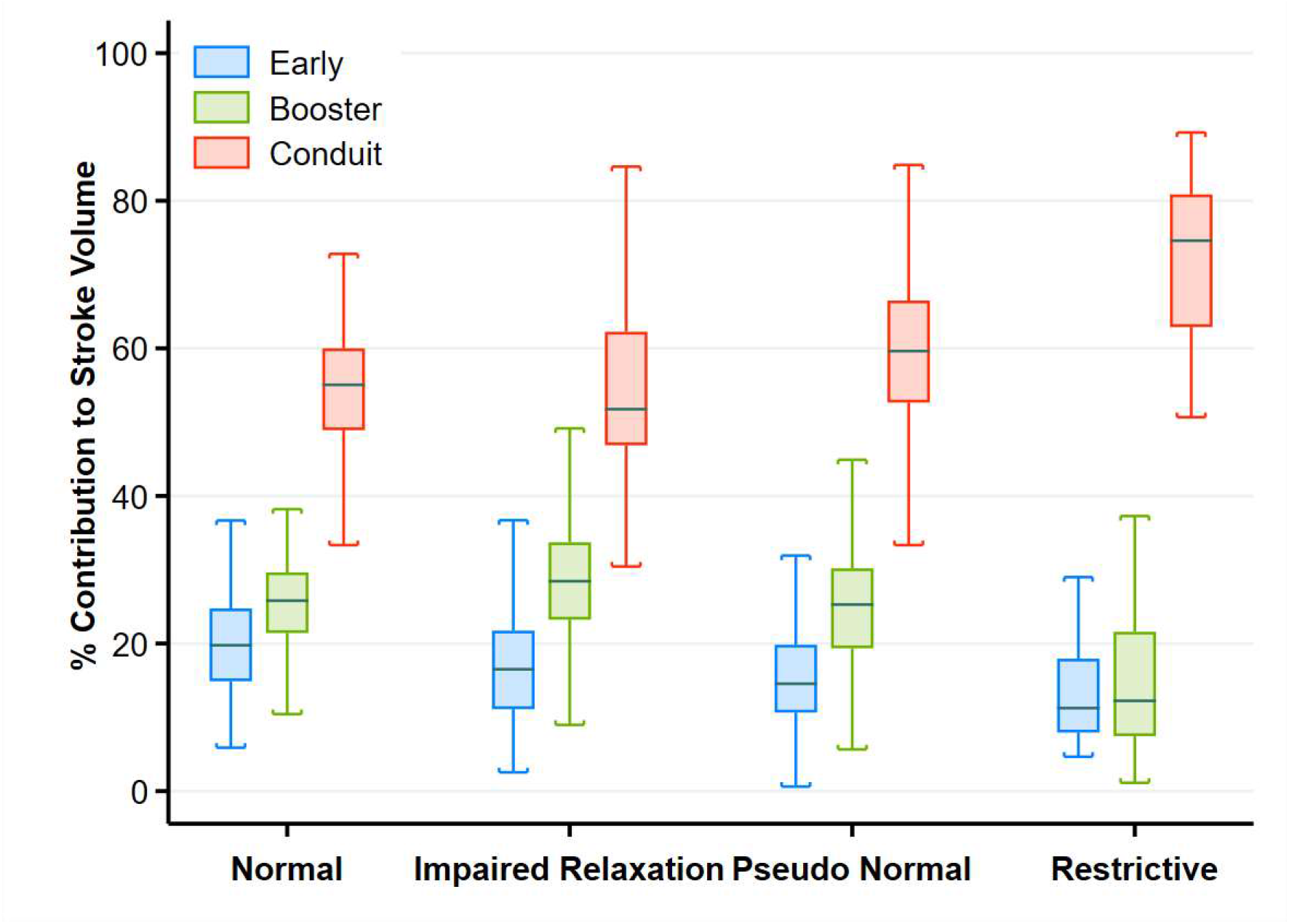
Figure 1: Box-and-whisker plot and scatter plot of the 3 components of left ventricular filling at various grades of diastolic function. The line within the box denotes the median and the box spans the interquartile range (25–75th percentiles). Whiskers extend from the 5^th^ to 95^th^ percentiles. (Kruskal-Wallis test, *P*<0.001 for all comparisons)

In subjects with normal diastolic function, the median contribution of LA conduit volume to LV filling was 55% [IQR 49–60%]. The conduit volume increases in grade II diastolic dysfunction (median 60%, IQR 53–66%; *P*<0.001 vs. normal diastolic function), thus compensating for the reduction of the early passive filling and further increases in grade III diastolic dysfunction (median 75%, IQR 63–81%; *P*<0.001 vs. pseudonormal diastolic function) to compensate for, or attenuate, the marked reduction in both early passive filling and booster function, thus maintaining ventricular filling and stroke volume (Central Illustration, A). Similar patterns were observed in the subgroup of patients with preserved LVEF (supplemental figure 1A) and mid-range or reduced LVEF (supplemental figure 1B) and when expressed as absolute volumes (supplemental figure 2). Overall, in advanced diastolic dysfunction, LV filling occurred predominantly from the pulmonary veins rather than from blood that is stored in the LA at end systole (Central Illustration, **B**).

Regardless of diastolic function, there was a strong inverse relationship between the contribution of early passive and booster volumes and the conduit volume for every level of SV (Supplemental Figure 3).

The conduit flow rate into the LV was calculated in a subset of 57 patient and followed a nonlinear pattern with a rapid flow at the beginning of diastole and near-plateau at the end of diastole. There was a progressive increase in the slope of the steep portion of the conduit flow curve with worsening diastolic function (Central illustration **C**). Compared with early diastole conduit flow rate in patients in the normal diastolic function (n=15, 133 mL/sec, 95% CI 118– 147, *P*=0.22) the conduit flow rate slope increased to 147 mL/sec [95% CI 129–165], 159 mL/sec [95% CI 144–173, *P*=0.01], and 234 mL/sec [95% CI 220–253, *P*=0.<0.001] in patients with inpaired relaxation, pseudonormal and restrictive filling, respectively.

### Association between conduit volume and pulmonary hypertension

Augmentation of conduit volume requires an increase in pulmonary venous pressure. Therefore, we tested the association between conduit volume contribution to SV and echocardiographic PASP. Conduit volume contributed 71% [IQR 60 to 81%] and 56% [IQR 50 to 63%] to left ventricular SV in patients with PAP above and below 50 mm Hg, respectively. Using PASP as a continuous variable in a cubic spline regression demonstrated a biphasic relationship between conduit function and PASP with an initial flat slope followed by an increase in a near-linear fashion when conduit contribution to SV increases above 60% (Figure 2A). Logistic regression demonstrated that the probability of PASP≥50 mm Hg increased with increasing conduit contribution to SV (Figure 2B).

**Figure 2:**
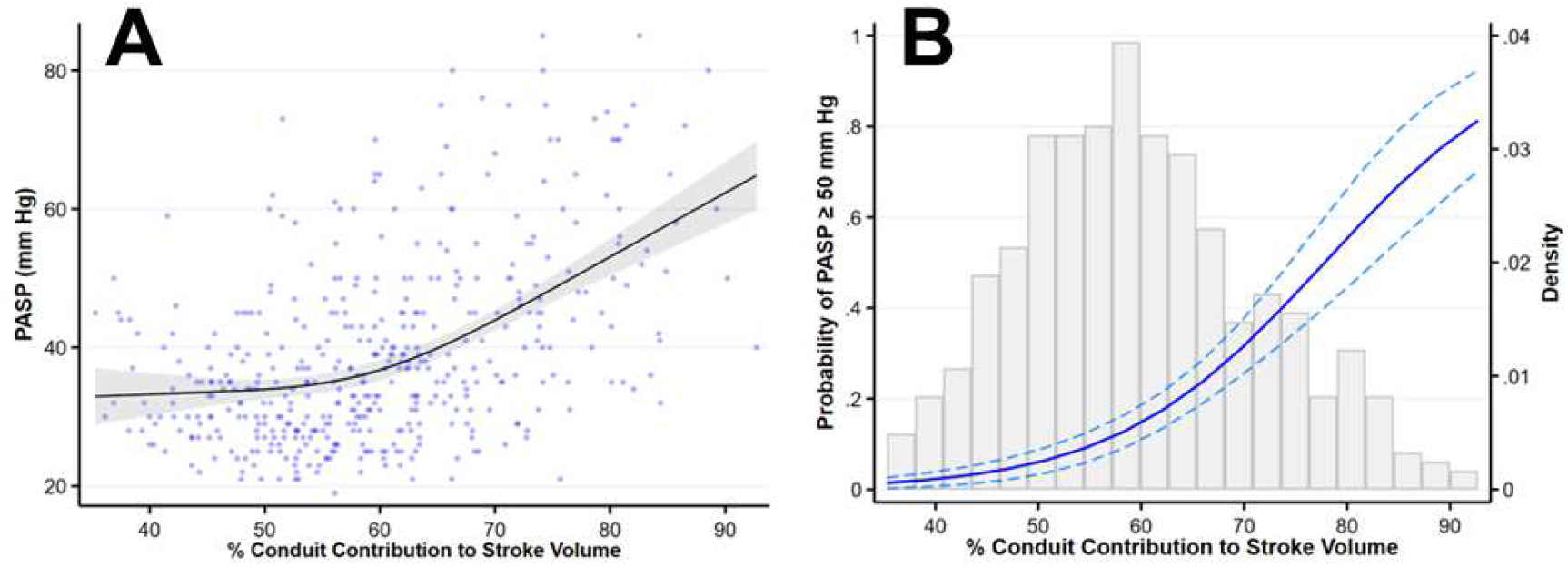
Relationship between conduit volume and pulmonary artery pressure. (A) Fractional polynomial regression of the relationship between PAP and percent contribution of conduit volume to left ventricular stroke volume. (B) Probability of PAP ≥50 mm Hg according to percent contribution of conduit volume to left ventricular stroke volume.

### Effect of LA remodeling on LA functions

We tested the effect of LA remodeling (demonstrated by increased LA volume) on each of the components of LA function. There was an inverted U-shaped relationship between LA volume and LA reservoir volume (*P* for quadratic trend <0.001), such that LA reservoir volume increases with mild-moderate increase in LAESVI but decreases with severe LA enlargement (Figure 3A). LA expansion index (reservoir function) was inversely related to LA enlargement (Figure 3B). LAEF**_passive_** also decreased with increasing LAVI (Figure 3C). Although LA enlargement was associated with a progressive increase in LA volume pre-A wave (*P*<0.001; Figure 3D), the LAEF**_contractile_** progressively decreased (Figure 3E), resulting in an impaired compensation for the reduced LA passive filling. By contrast, the conduit volume increased with increasing LA volume (Figure 3F).

**Figure 3:**
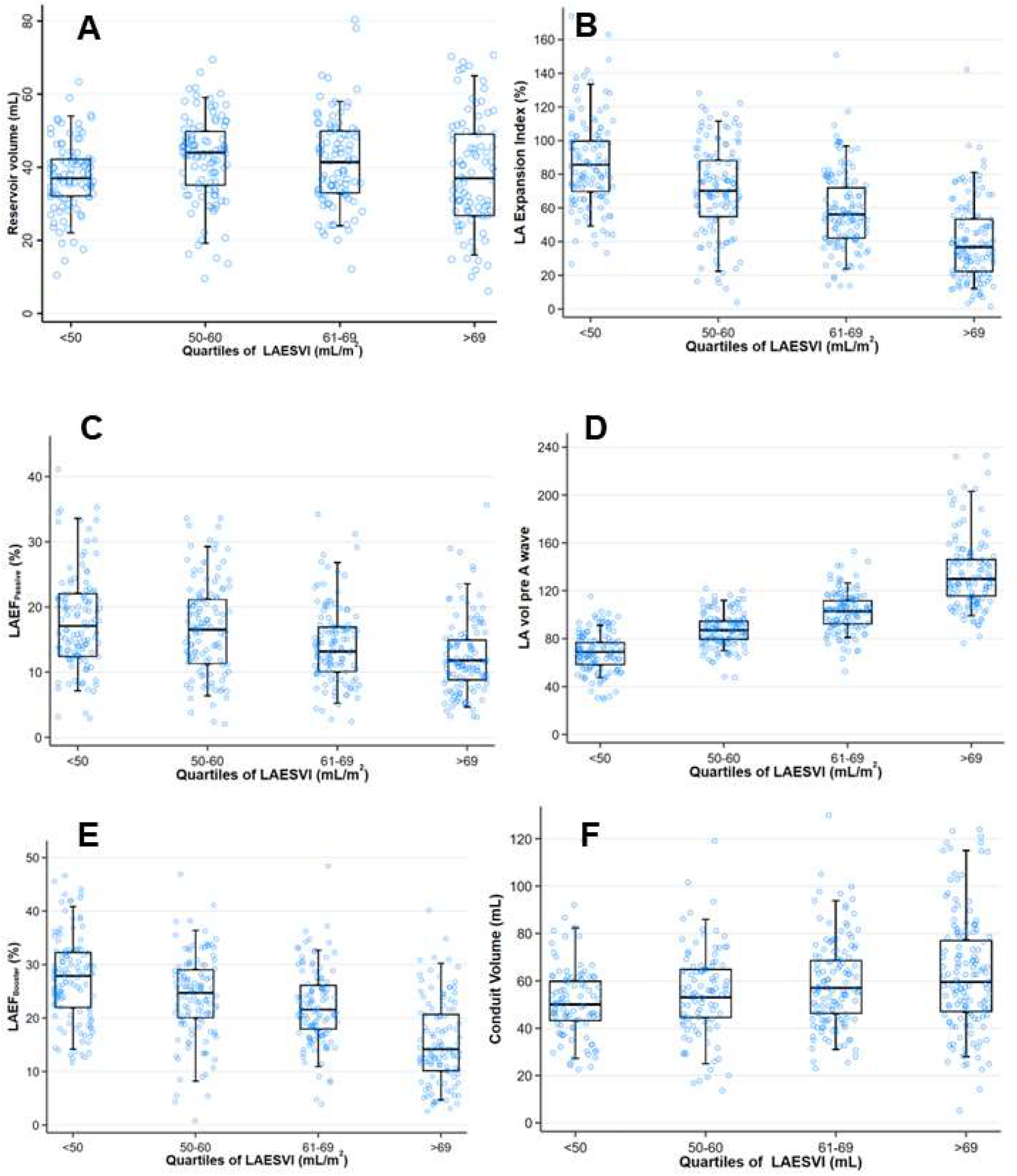
Effect of left atrial remodeling on the components of left atrial function. **A.** U-shaped relationship between LA volume and LA reservoir volume; **B.** LA expansion index; **C.** LA emptying fraction during passive LV filling; **D.** LA pre-A volume; **E.** LA emptying fraction during active atrial systole; **F.** Conduit volume.

### Early diastolic mitral inflow velocity and Conduit flow

The echocardiographic early diastolic mitral inflow velocity was not significantly associated with early passive filling volume (Figure 4A) and flow rate (Figure 4B). However, there was a moderate association between early diastolic mitral inflow velocity and conduit volume (Figure 4C) and a stronger correlation with maximal conduit flow rate (Figure 4D). Figure 5 depicts the early passive, conduit and atrial systolic maximal flow rates according to the echocardiographic diastolic grade, demonstrating that the conduit maximal flow rate increases with worsening LVDD whereas the maximal flow rate of early passive filling decreases. Overall, these data indicates that the increase of mitral E-wave velocity with worsening LVDD is related to the concomitant increase in conduit flow rather than the early passive filling.

**Figure 4:**
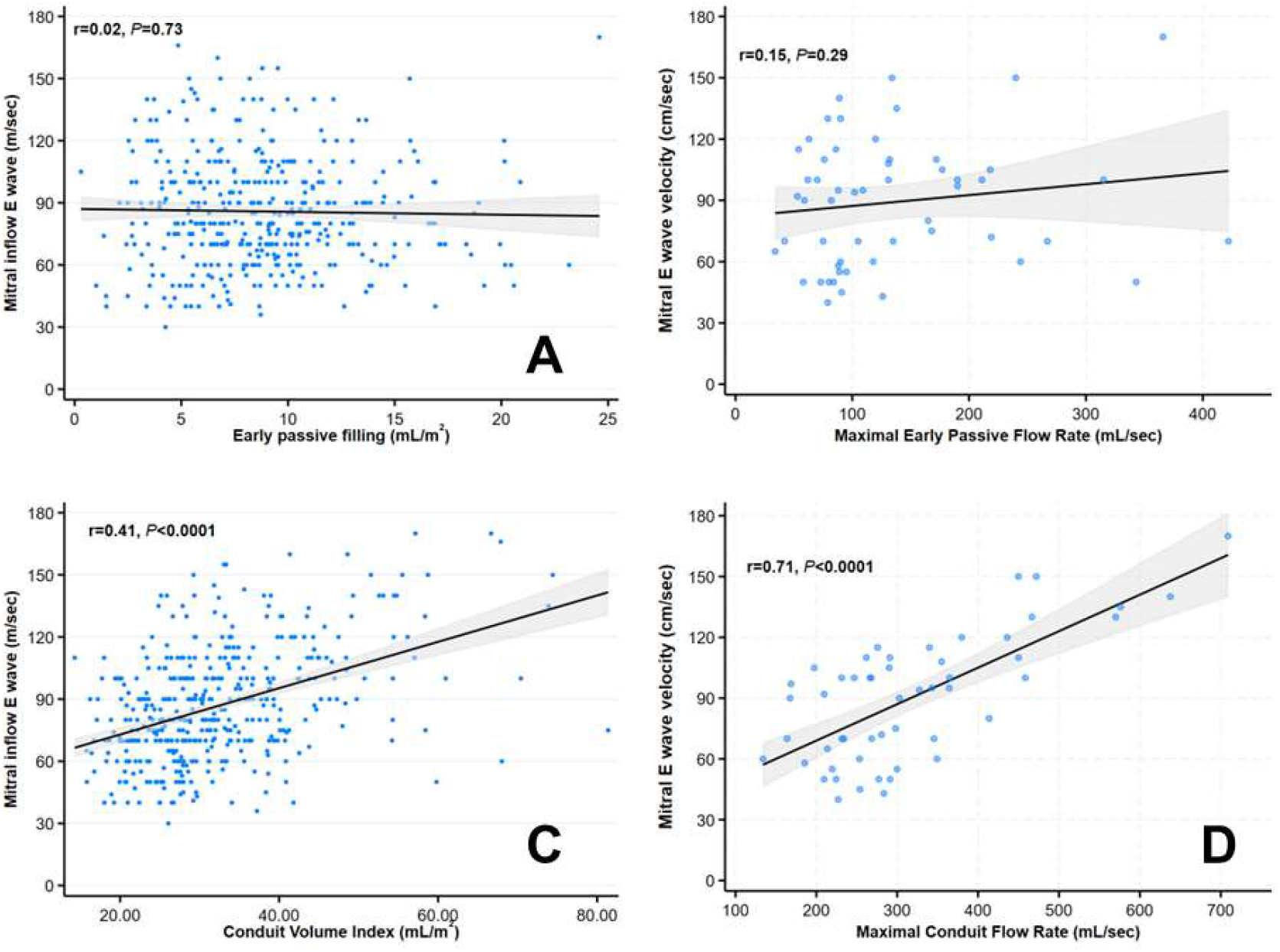
Relationship between peak early diastolic mitral inflow velocity by Doppler and **(A)** Early passive LA emptying volume **(B)** Maximal early passive flow rate **(C)** Conduit volume **(D)** Maximal conduit flow rate.

**Figure 5:**
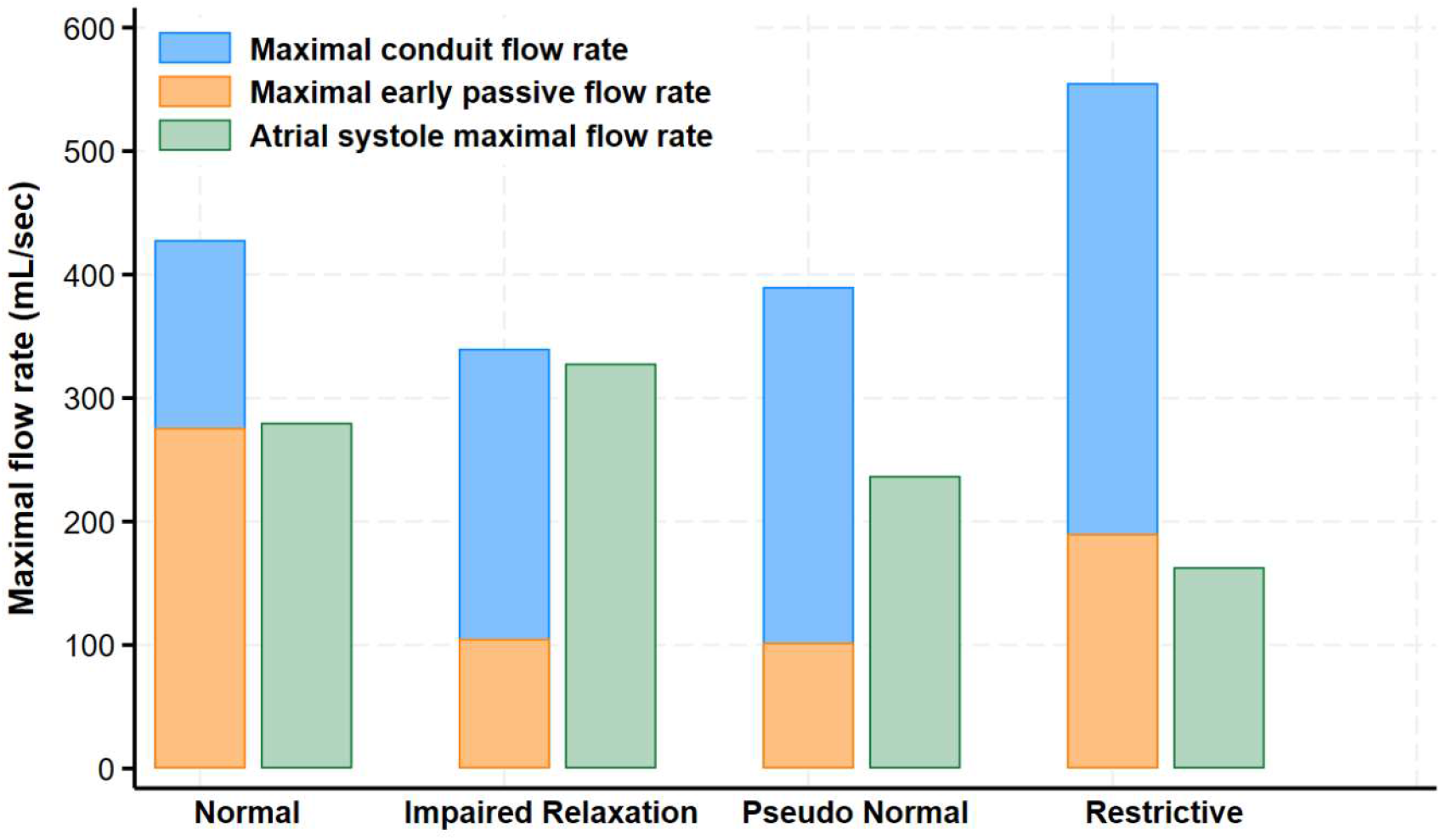
Median values of early passive, conduit, and atrial systolic maximal flow rates according to the echocardiographic diastolic grade. The early passive and conduit flow occur nearly simultaneously and therefore represented on the same bar graph with their relative contribution to the mitral E wave.

### Conduit function in aortic stenosis

In the prospective group of AS patients, advanced diastolic dysfunction was common (81% with grade II or III). We observed the same pattern of phasic changes in LA volumes as in the retrospective cohort (Figure 6). Worsening diastolic function was associated with a progressive reduction in early passive filling and booster function and with a progressive increase in conduit function (median contribution to LV filling 58% and 64% in grade II and grade III diastolic dysfunction, respectively).

**Figure 6:**
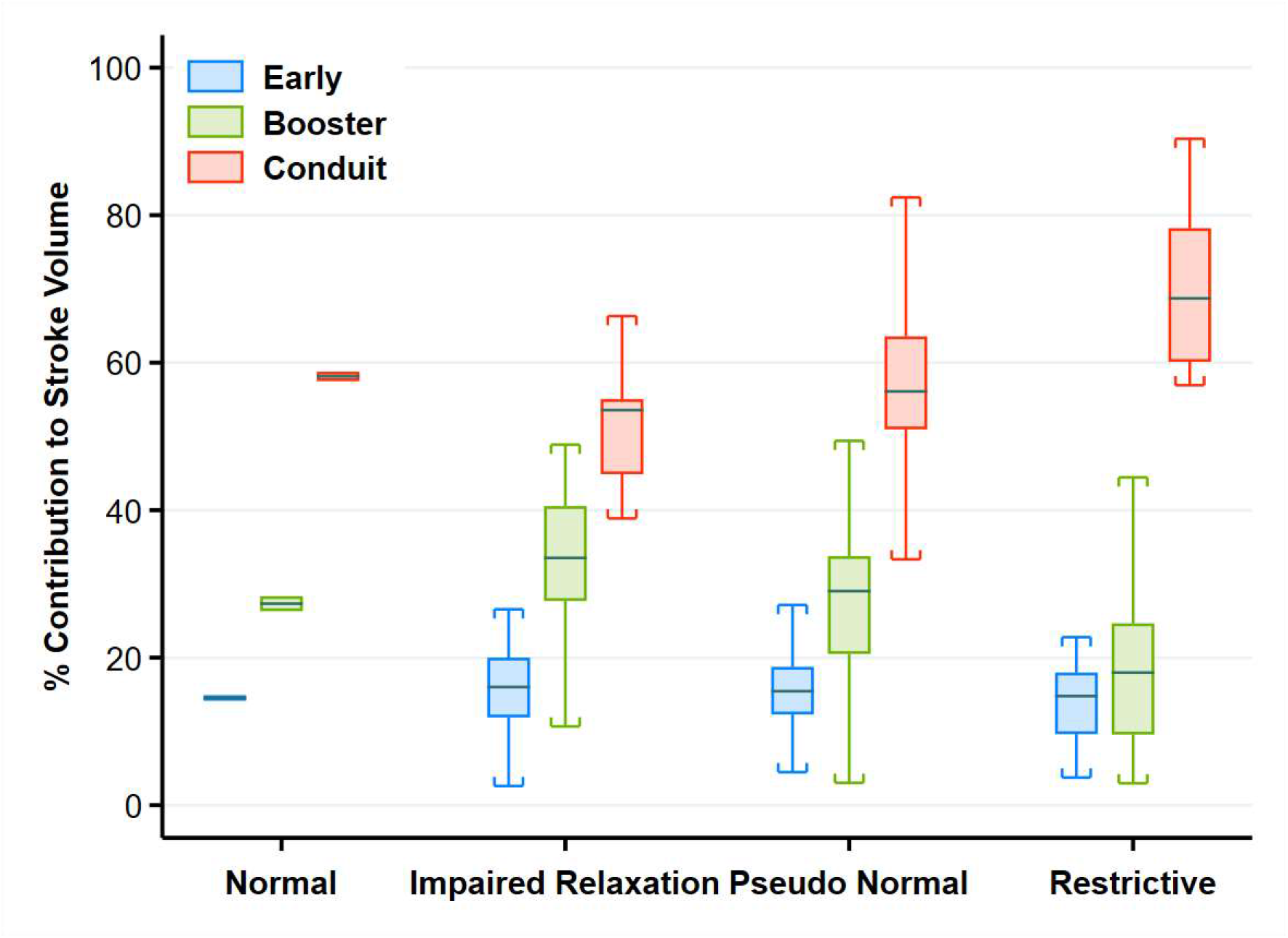
Box-and-whisker plot and scatter plot of the 3 components of left ventricular filling at various grades of diastolic function in patients with severe aortic stenosis.

## Discussion

We measured CTCA-derived LA phasic volumes as absolute values and in terms of their contribution to LV filling as well as maximal flow rates. Volumetric analysis of LA function provided a simple and intuitive interpretation of LA function across the spectrum of LV diastolic dysfunction. Progressive diastolic dysfunction was initially associated with a reduction of early passive emptying volume, accompanied by a compensatory increase in active emptying, followed by (in grade 2 dysfunction) failure of the booster function (active emptying) to compensate for the reduction in early emptying. In advanced diastolic dysfunction, the current study establishes the crucial role of augmented conduit volume as the major compensatory mechanism to maintain adequate LV filling that mitigates the reductions in both passive and booster functions. The increase in conduit volume and flow despite progressive diastolic dysfunction and increasing LV diastolic pressures involves an increase in pulmonary venous pressure.

It has been previously estimated that conduit function constitutes approximately 20% to 30% of the SV (24). In the current analysis we found that even with normal diastolic function, the conduit volume contributes ∼50% to SV and can increase to more than 70% in severe LVDD. Therefore, in advanced diastolic dysfunction, LV filling occurs predominantly from the pulmonary veins rather than from blood that is stored in the LA at end systole.

### Conduit flow and the echocardiographic mitral E velocity

LV filling due to the early passive LA emptying and conduit flow occur simultaneously in early diastole, both affecting the echocardiographic mitral E velocity. Importantly, the early passive volume and flow rate progressively decrease with worsening diastolic function and cannot explain the increasing mitral E wave velocity. The conduit flow, however, continues to increase with worsening diastolic function and accounts for most of the mitral E wave velocity.

### Mechanisms of compensation for increased LV diastolic pressures

Maintaining SV and cardiac output to supply the tissues is fundamental to circulatory physiology. With mild diastolic dysfunction, the atrial booster function can compensate for the reduction in early passive filling. At the end of diastasis, the LA volume remains elevated, and the booster function increases via the Frank-Starling mechanism. This represents the initial compensatory process for the early stage of LV diastolic dysfunction.

Over time, atrial myocyte loss and atrial fibrosis progressively diminish the ability of the booster pump function to compensate for the reduced early passive LA function (15). Notably, the booster fraction may still be in the “normal” range but not sufficient to compensate for the progressive reduction in early ventricular filling. At this point, a second compensatory mechanism, namely an increase in conduit volume, becomes the principal instrument to preserve SV and systemic flow.

The present study also demonstrates that atrial remodeling is often maladaptive with respect to the early passive and booster functions. Increasing LA volume was associated with a reduction in LA expansion index (a measure of reservoir function) as well as both LAEF**_passive_** and LAEF**_active_**. By contrast, the conduit volume is an effective compensatory mechanism to maintain normal SV that is largely independent of LA remodeling and function. The conduit volume can therefore offset the reductions in both early filling and booster function and can serve as an effective compensatory mechanism to maintain adequate LV filling.

### Pulmonary hypertension and conduit volume

LA conduit function is believed to be mainly determined by the rate of left ventricular relaxation as blood is drawn into the LV via the resulting suction (7,25,26). With worsening of LV diastolic function, however, such a mechanism cannot explain the observed increase in conduit volume (with the concomitant decrease in early passive LV filling).

Increasing conduit volume in the face of increasing LV diastolic pressures requires further increase in pulmonary venous pressure. In normal subjects, Bowman and Kovacs (7) have previously demonstrated a positive relationship between the LA conduit volume and diastolic pulmonary venous flow (pulmonary D wave), that is known to be augmented with moderate or severe diastolic dysfunction (27).The increase in pulmonary venous pressure can be attributed to the transpulmonary propagation of pulmonary artery pressure, which closely correlates with pulmonary venous pressure over a wide range of loading conditions (28). Therefore, the present study strongly supports the concept that the conduit volume is augmented by an increase in pulmonary venous pressure in patients with LV diastolic dysfunction. Importantly, in the presence of an adequate compensatory booster function, an increase in pulmonary venous pressure is not required to maintain adequate LV filling.

### Discrepancy with previous studies

Multiple studies reported a reduction in conduit function (in conjunction to a reduction in reservoir and booster functions) with aging and in both HFpEF and HFrEF (9–11,17,29). A recent meta-analysis of 22 studies comprising of 1974 patient with HFpEF, reported a reduction in LV reservoir, booster, and conduit functions (30). However, a decrease in the conduit volume in advanced diastolic dysfunction implies a reduction of all 3 components contributing to LV filling, inevitably leading to a marked reduction in resting LV filling and hence in SV.

Estimation of conduit function using only LA deformation imaging does not adequately capture the conduit volume during diastole. For example, conduit function as assessed by deformation analysis (2) follows the reduction in LA volume during early filling and corresponds only to passive LA emptying in the volumetric CTCA analysis. Therefore, this measure is expected to decline with worsening diastolic function. However, because conduit flow involves the transfer of blood into the left ventricle without affecting LA volume, it cannot be measured by deformation imaging of the LA. Therefore, the discrepancy between the current study and previous echocardiographic studies (10,11,17,29,30) is likely driven from the way passive, active and conduit functions are captured and estimated.

### Conduit volume in aortic stenosis

In patients with severe AS, the development of symptoms is associated with impaired diastolic function and LA dilatation independent of stenosis severity (31). In these patients, LA booster-pump function has a crucial role in counteracting the increased LV end-diastolic pressure, thus maintaining an adequate end-diastolic volume and sustaining cardiac output (32). Loss of “atrial kick” is often a concern in severe AS. However, Kroetz et al. reported that loss of effective atrial systole had little influence on steady state or integrated cardiac performance in patients with severe AS, concluding that when the contribution of atrial contraction is lost, other adaptive mechanisms partially or completely compensate (33). In the current study, LA booster-pump function was impaired in many patients (who were symptomatic and candidates for valve intervention) making the conduit volume the dominant contributor to LV filling in these patients.

### Study limitations

It is important to consider several limitations pertinent to the methods of this study. First, this was a single-center post-hoc analysis of our CCTA data, and thus, the results must be regarded as hypothesis generating and exploratory and require validation in other studies. Sampling bias may have occurred because patients at higher risk were more likely to be referred for CCTA. In addition, there is a bias generated by the study inclusion criteria requiring spiral CT studies as well as a recent echocardiogram. As this was a cross-sectional study, we could only observe patients at a single timepoint.

## Conclusion

An increase in conduit volume emerged as a major compensatory mechanism to maintain LV filling in advanced diastolic dysfunction when LA booster function fails. The increase in conduit volume despite progressive diastolic dysfunction and increasing LV diastolic pressures is accomplished by an increase in pulmonary pressure.

## Data Availability

All data produced in the present study are available upon reasonable request to the authors

---

**Central Illustration:**
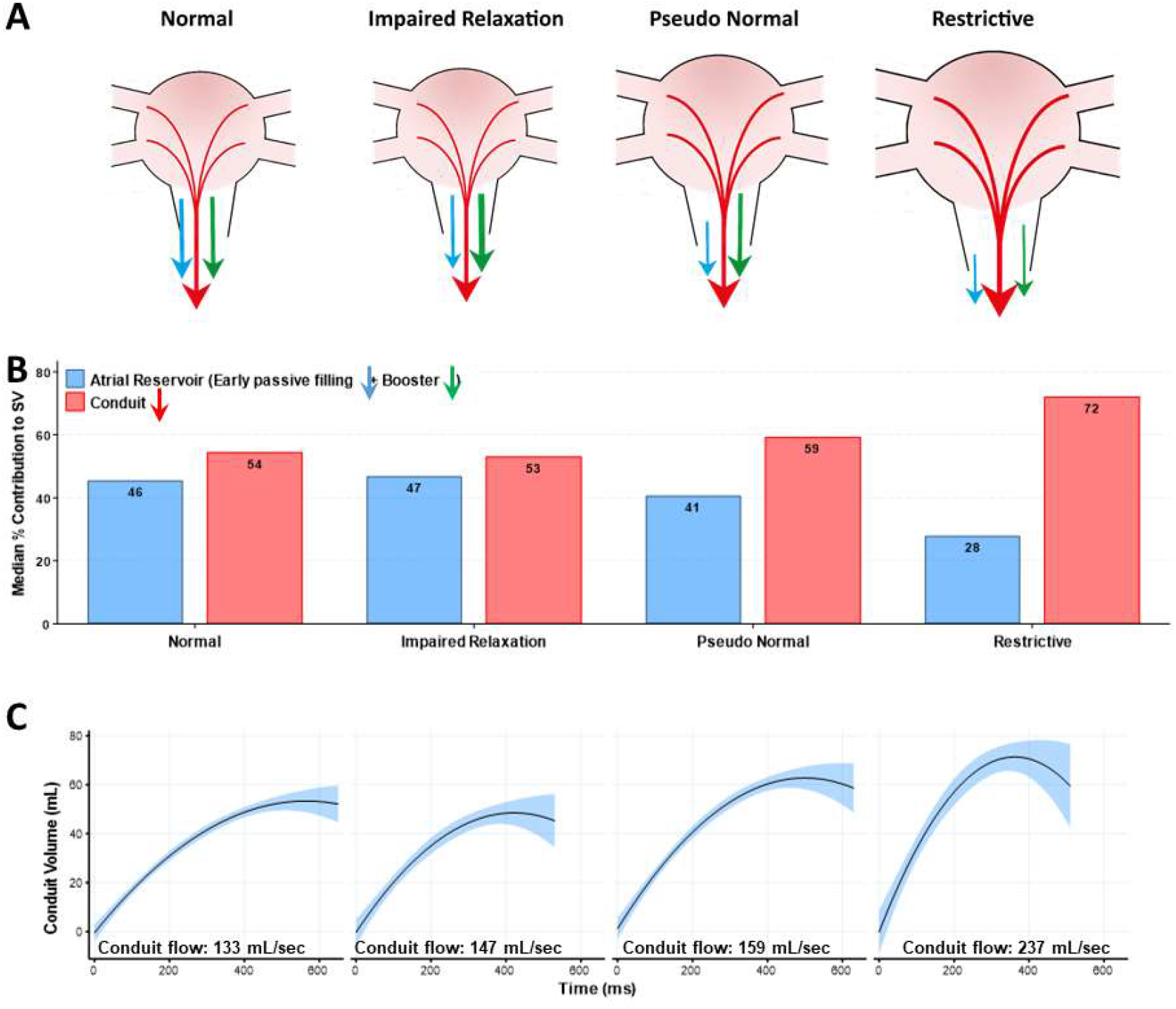
Progression of diastolic dysfunction: (A) With impaired relaxation, the atrial systole compensated for the reduction in early passive filling. With more advanced diastolic dysfunction, the booster pump function declines. At this stage, an increase in conduit flow becomes the principal mechanism to compensate for the reductions in both early passive and booster functions and contributes over 70% of LV filling. **(B)** Therefore, in advanced diastolic dysfunction, LV filling occurs predominantly from the pulmonary veins rather than from blood stored in the LA. **(C)** The conduit flow rate increases progressively with the worsening of diastolic function and accounts for most of the increased mitral E wave velocity in advanced diastolic dysfunction.

## References

1. Rossi A, Gheorghiade M, Triposkiadis F, Solomon SD, Pieske B, Butler J. Left atrium in heart failure with preserved ejection fraction: structure, function, and significance. Circ Heart Fail 2014;7:1042–9.

2. Hoit BD. Left atrial size and function: role in prognosis. J Am Coll Cardiol 2014;63:493–505.

3. Nishimura RA, Tajik AJ. Evaluation of Diastolic Filling of Left Ventricle in Health and Disease: Doppler Echocardiography Is the Clinician’s Rosetta Stone. Journal of the American College of Cardiology 1997;30:8–18.

4. Welles CC, Ku IA, Kwan DM, Whooley MA, Schiller NB, Turakhia MP. Left atrial function predicts heart failure hospitalization in subjects with preserved ejection fraction and coronary heart disease: longitudinal data from the Heart and Soul Study. J Am Coll Cardiol 2012;59:673–80.

5. Melenovsky V, Borlaug BA, Rosen B, et al. Cardiovascular features of heart failure with preserved ejection fraction versus nonfailing hypertensive left ventricular hypertrophy in the urban Baltimore community: the role of atrial remodeling/dysfunction. J Am Coll Cardiol 2007;49:198–207.

6. Pagel PS, Kehl F, Gare M, Hettrick DA, Kersten JR, Warltier DC. Mechanical function of the left atrium: new insights based on analysis of pressure-volume relations and Doppler echocardiography. Anesthesiology 2003;98:975–94.

7. Bowman AW, Kovacs SJ. Left atrial conduit volume is generated by deviation from the constant-volume state of the left heart: a combined MRI-echocardiographic study. Am J Physiol Heart Circ Physiol 2004;286:H2416–24.

8. Nagueh SF, Smiseth OA, Appleton CP, et al. Recommendations for the Evaluation of Left Ventricular Diastolic Function by Echocardiography: An Update from the American Society of Echocardiography and the European Association of Cardiovascular Imaging. Eur Heart J Cardiovasc Imaging 2016;17:1321–1360.

9. Kusunose K, Motoki H, Popovic ZB, Thomas JD, Klein AL, Marwick TH. Independent association of left atrial function with exercise capacity in patients with preserved ejection fraction. Heart 2012;98:1311–7.

10. Otani K, Takeuchi M, Kaku K, et al. Impact of diastolic dysfunction grade on left atrial mechanics assessed by two-dimensional speckle tracking echocardiography. J Am Soc Echocardiogr 2010;23:961–7.

11. Okamatsu K, Takeuchi M, Nakai H, et al. Effects of aging on left atrial function assessed by two-dimensional speckle tracking echocardiography. J Am Soc Echocardiogr 2009;22:70–5.

12. Yamano M, Yamano T, Iwamura Y, et al. Impact of Left Ventricular Diastolic Property on Left Atrial Function from Simultaneous Left Atrial and Ventricular Three-Dimensional Echocardiographic Volume Measurement. Am J Cardiol 2017;119:1687–1693.

13. Marino PN, Degiovanni A, Zanaboni J. Complex interaction between the atrium and the ventricular filling process: the role of conduit. Open Heart 2019;6:e001042.

14. Triposkiadis F, Pieske B, Butler J, et al. Global left atrial failure in heart failure. Eur J Heart Fail 2016;18:1307–1320.

15. Thomas L, Marwick TH, Popescu BA, Donal E, Badano LP. Left Atrial Structure and Function, and Left Ventricular Diastolic Dysfunction: JACC State-of-the-Art Review. J Am Coll Cardiol 2019;73:1961–1977.

16. Ghersin I, Ghersin E, Abadi S, et al. Assessment of Diastolic Function in Hypertrophic Cardiomyopathy by Computed Tomography-Derived Analysis of Left Ventricular Filling. J Comput Assist Tomogr 2017;41:339–343.

17. Melenovsky V, Hwang SJ, Redfield MM, Zakeri R, Lin G, Borlaug BA. Left atrial remodeling and function in advanced heart failure with preserved or reduced ejection fraction. Circ Heart Fail 2015;8:295–303.

18. Russo C, Jin Z, Homma S, et al. LA Phasic Volumes and Reservoir Function in the Elderly by Real-Time 3D Echocardiography: Normal Values, Prognostic Significance, and Clinical Correlates. JACC Cardiovasc Imaging 2017;10:976–985.

19. Stefanadis C, Dernellis J, Toutouzas P. A clinical appraisal of left atrial function. Eur Heart J 2001;22:22–36.

20. Posina K, McLaughlin J, Rhee P, et al. Relationship of phasic left atrial volume and emptying function to left ventricular filling pressure: a cardiovascular magnetic resonance study. J Cardiovasc Magn Reson 2013;15:99.

21. Lessick J, Mutlak D, Efraim R, et al. Comparison Between Echocardiography and Cardiac Computed Tomography in the Evaluation of Diastolic Dysfunction and Prediction of Heart Failure. Am J Cardiol 2022;181:71–78.

22. Lessick J, Mutlak D, Mutlak M, et al. Left atrial function by cardiac computed tomography is a predictor of heart failure and cardiovascular death. Eur Radiol 2022;32:132–142.

23. Schweitzer A, Agmon Y, Aronson D, et al. Assessment of left sided filling dynamics in diastolic dysfunction using cardiac computed tomography. Eur J Radiol 2015;84:1930–7.

24. Bisbal F, Baranchuk A, Braunwald E, Bayes de Luna A, Bayes-Genis A. Atrial Failure as a Clinical Entity: JACC Review Topic of the Week. J Am Coll Cardiol 2020;75:222–232.

25. Nikitin NP, Witte KK, Thackray SD, Goodge LJ, Clark AL, Cleland JG. Effect of age and sex on left atrial morphology and function. Eur J Echocardiogr 2003;4:36–42.

26. Hoit BD. Left Atrial Reservoir Strain: Its Time Has Come. JACC Cardiovasc Imaging 2022;15:392–394.

27. Oh JK, Appleton CP, Hatle LK, Nishimura RA, Seward JB, Tajik AJ. The noninvasive assessment of left ventricular diastolic function with two-dimensional and Doppler echocardiography. J Am Soc Echocardiogr 1997;10:246–70.

28. Chaliki HP, Hurrell DG, Nishimura RA, Reinke RA, Appleton CP. Pulmonary venous pressure: relationship to pulmonary artery, pulmonary wedge, and left atrial pressure in normal, lightly sedated dogs. Catheter Cardiovasc Interv 2002;56:432–8.

29. von Roeder M, Rommel KP, Kowallick JT, et al. Influence of Left Atrial Function on Exercise Capacity and Left Ventricular Function in Patients With Heart Failure and Preserved Ejection Fraction. Circ Cardiovasc Imaging 2017;10.

30. Khan MS, Memon MM, Murad MH, et al. Left atrial function in heart failure with preserved ejection fraction: a systematic review and meta-analysis. Eur J Heart Fail 2020;22:472–485.

31. Dahl JS, Christensen NL, Videbaek L, et al. Left ventricular diastolic function is associated with symptom status in severe aortic valve stenosis. Circ Cardiovasc Imaging 2014;7:142–8.

32. Carabello BA. How does the heart respond to aortic stenosis: let me count the ways. Circ Cardiovasc Imaging 2013;6:858–60.

33. Kroetz FW, Leonard JJ, Shaver JA, Leon DF, Lancaster JF, Beamer VL. The effect of atrial contraction on left ventricular performance in valvular aortic stenosis. Circulation 1967;35:852–67.

