## Supplemental Data for "Enhanced Conduit Flow Compensates for the Reduction in Left Atrial Passive and Booster Functions in Advanced Diastolic Dysfunction"

**SUPPLEMENTAL MATERIAL**

**Supplemental Figure 1A**

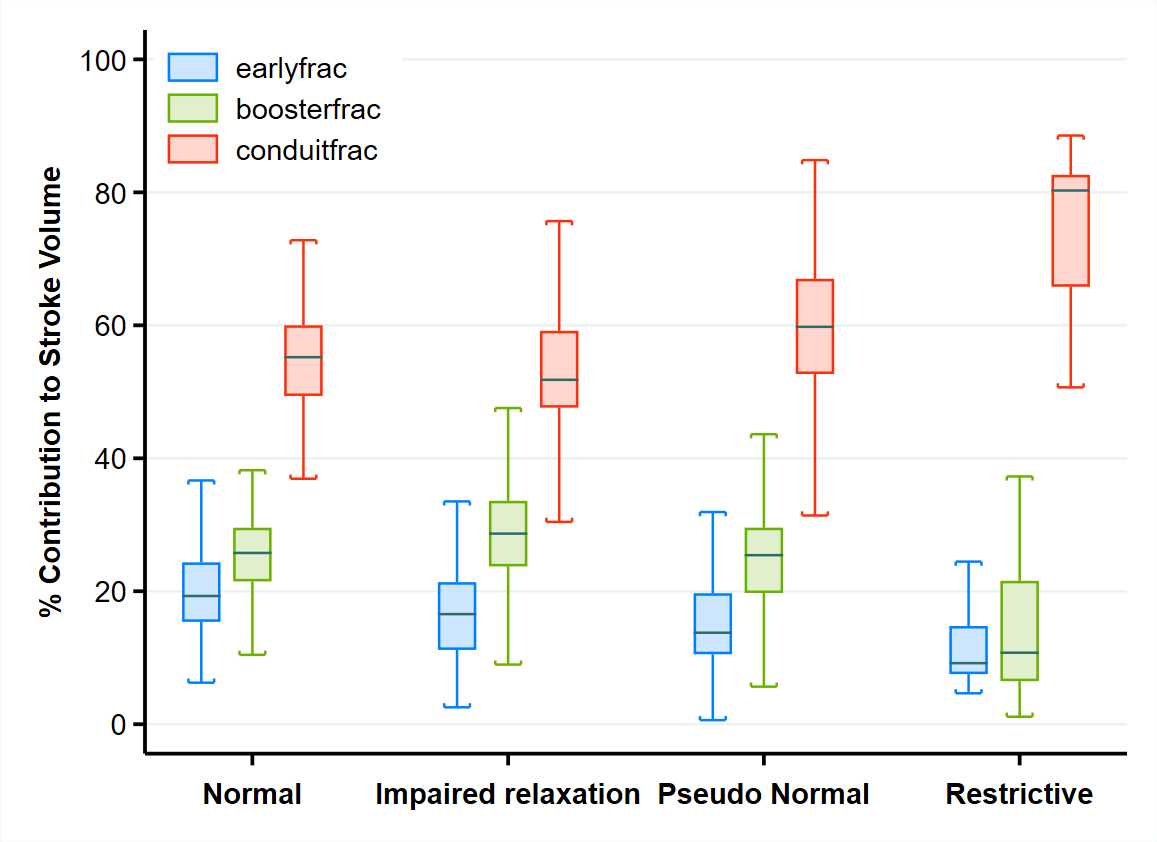

**Supplemental Figure 1A:** Box-and-whisker plot and scatter plot of the 3 components of left ventricular filling at various grades of diastolic function for patient with preserved ejection fraction (Left ventricular ejection fraction ≥50%).

**Supplemental Figure 1B**

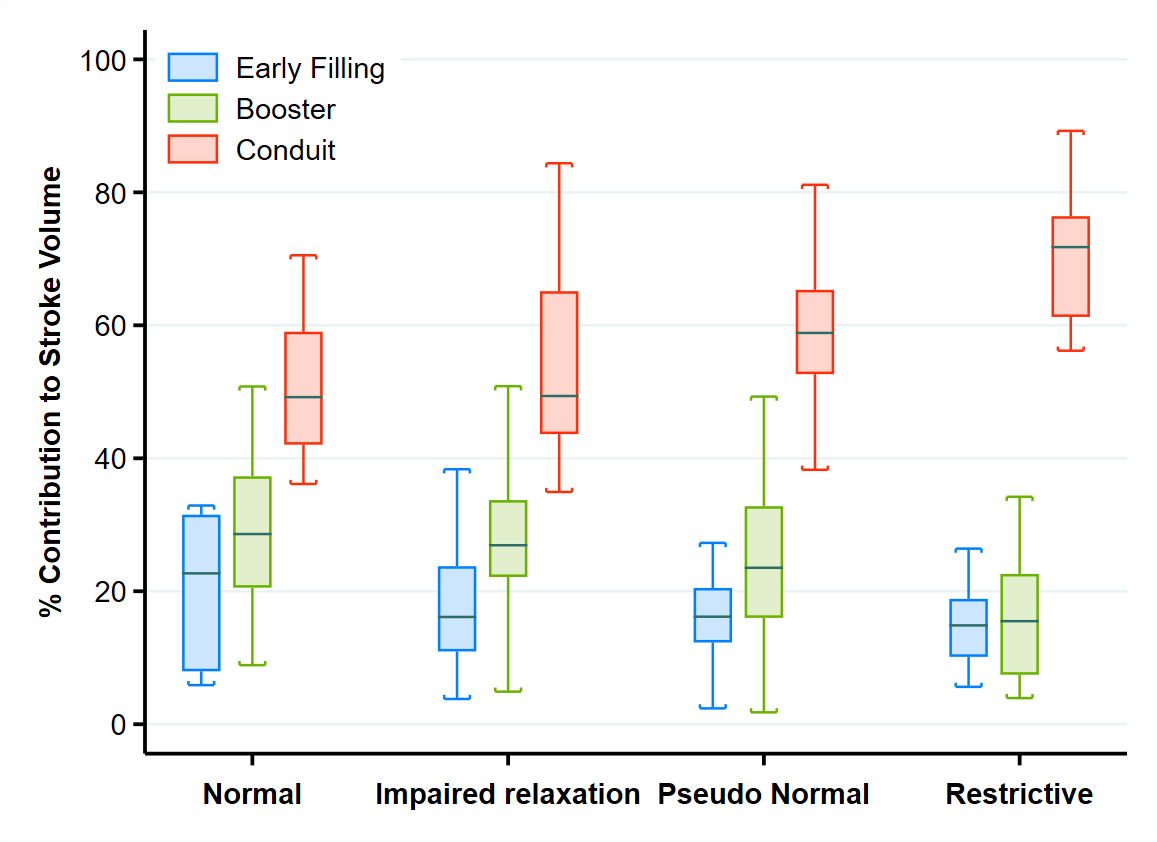

**Supplemental Figure 1B:** Box-and-whisker plot and scatter plot of the 3 components of left ventricular filling at various grades of diastolic function for patient with mid-range and reduced ejection fraction (Left ventricular ejection fraction <50%).

**Supplemental Figure 2**

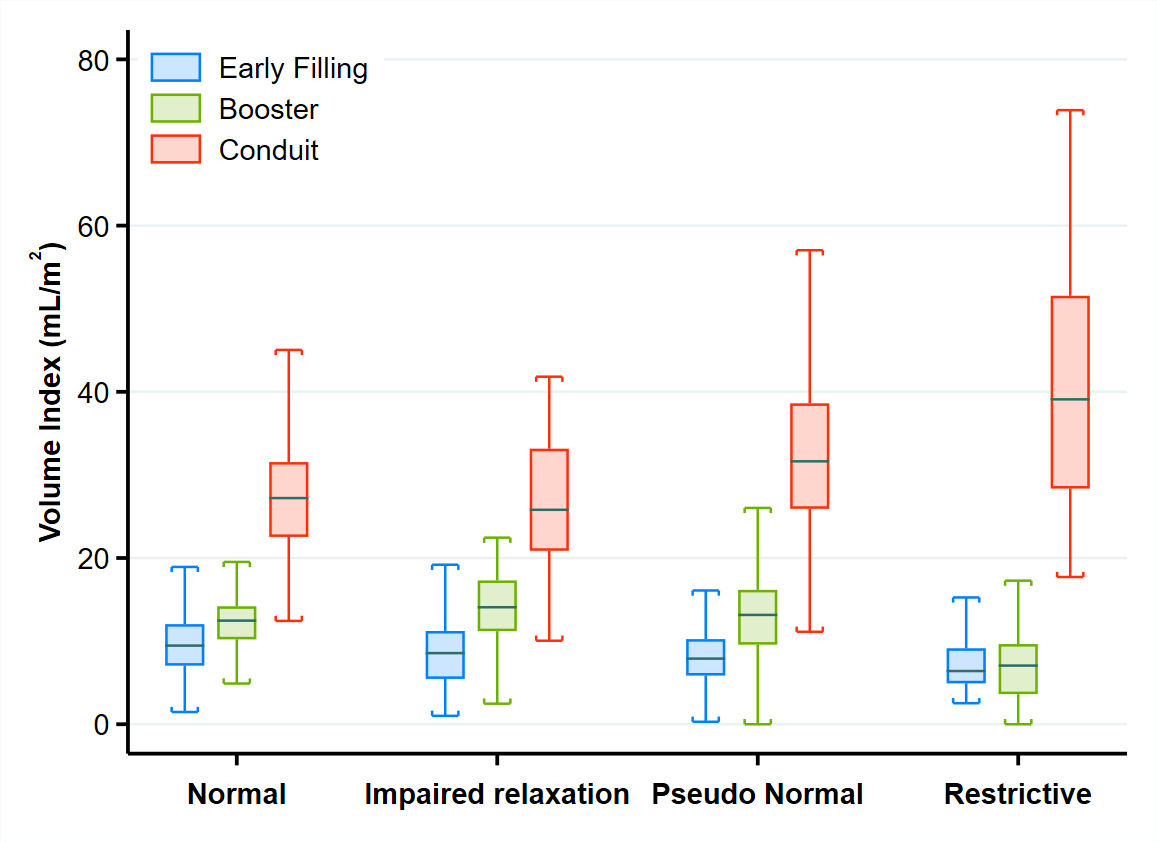

**Supplemental Figure 2:** Box-and-whisker plot and scatter plot of the 3 components of left ventricular filling at various grades of diastolic function expressed as absolute volumes.

**Supplemental Figure 3**

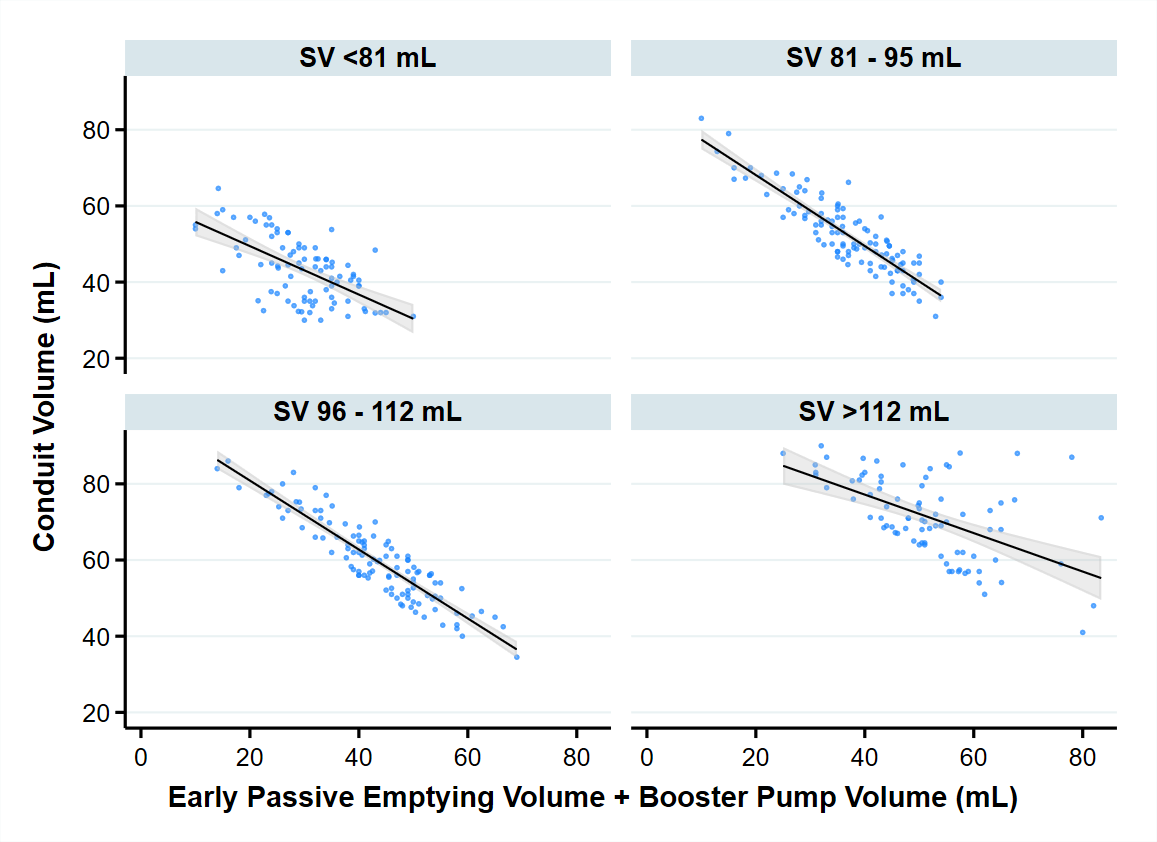

**Supplemental Figure 3:** Relationship of the combined early passive volume and booster pump volume with the conduit volume, depicted across quartiles of stroke volume (SV). For every level of SV, the conduit volume is inversely related to the sum of early passive volume and booster pump volume, thus preventing a reduction in SV.
